# Practical Management of Adverse Events Associated with Bispecific Antibodies for the Treatment of Multiple Myeloma: A Qualitative Interview Study

**DOI:** 10.64898/2026.04.24.26350878

**Authors:** Tisheeka R. Graham, Michael White, Brandon Blue, Monique Hartley-Brown, Bradley D. Hunter, Chanh Huynh, Nisha Joseph, Amany Keruakous, Darren Pan, Priya Rudolph, Rishi Sawhney, Attaya Suvannasankha

## Abstract

**PURPOSE:** Bispecific antibodies (BsAbs) represent a major advancement in the management of relapsed/refractory multiple myeloma (RRMM), offering high response rates even in heavily pretreated patients. However, their use presents operational, safety, and supportive care complexities that require coordinated care teams, and evolving infrastructure. This manuscript summarizes best practice recommendations for adverse event (AE) management, outpatient operational models, referral pathways, and emerging strategies to optimize long-term tolerability.

**METHODS:** Medlive—A PlatformQ Health Brand conducted qualitative interviews of academic and community-based clinicians. Discussions focused on BsAb implementation, patient selection and counseling, and AE management. Experts provided recommendations on team-based protocols, transitions of care, and inpatient versus outpatient considerations.

**RESULTS:** Ten hematologists/oncologists (academic n=4; community n=6) described practice patterns, barriers, and perspectives on BsAb use. BsAbs were consistently regarded as highly effective across multiple lines of therapy, particularly for patients without alternatives. Cytokine release syndrome (CRS) was the most common acute toxicity, generally low grade and managed effectively with early tocilizumab, including prophylactic use in outpatient settings. Immune effector cell-associated neurotoxicity syndrome (ICANS) was rare, mild, and best mitigated through early recognition and caregiver support. Infections, largely from BCMA-associated hypogammaglobulinemia, frequently interrupted therapy, necessitating antiviral prophylaxis, pneumocystis jirovecii pneumonia (PJP) prophylaxis, and intravenous immunoglobulin (IVIG). Outpatient step-up dosing is expanding, supported by prophylactic strategies and academic-community collaboration. Timely referral was emphasized to preserving eligibility. Major outpatient challenges included sequencing, infrastructure readiness, and standardized caregiver and staff education.

**CONCLUSION:** Effective community implementation of BsAbs requires multidisciplinary coordination, standardized AE protocols, infection prevention, and infrastructure to support monitoring, referrals, and equitable access. These measures are critical to ensure safe, sustainable integration of bispecific therapies and to optimize patient outcomes.

## Introduction

Despite recent advances in treatment, multiple myeloma (MM) is still considered an incurable malignancy.^1^ Even with aggressive first-line regimens (e.g., combinations of proteasome inhibitors, immunomodulatory agents, and anti-CD38 monoclonal antibodies) most patients inevitably relapse.^1,2^ Recent phase 3 data report 5-year overall survival rates exceeding 70% in trial populations;^3^ however, population-based estimates remain closer to 60%, underscoring the gap between clinical trial outcomes and real-world experience.^4^ Outcomes are particularly poor in patients with relapsed/refractory multiple myeloma (RRMM) who have become resistant to all three major drug classes,^5^ highlighting an urgent need for novel treatment approaches.

The pursuit for tolerable and effective treatments in the relapsed/refractory (RR) setting, especially for heavily treated patient populations, has resulted in novel therapies, including bispecific antibodies (BsAbs).^6–10^ By simultaneously targeting CD3-expressing T cells and tumor-associated antigens such as B-cell maturation antigen (BCMA) or G protein–coupled receptor class C group 5 member D (GPRC5D), BsAbs redirect host immunity toward malignant plasma cells.^11^ These agents have demonstrated high response rates, including in triple-class refractory patients, and are viewed by many clinicians as a transformative addition to the therapeutic arsenal.^12–15^ The introduction of BsAbs into real-world practice, however, presents several challenges. To ensure equitable access and optimal outcomes, these therapies must be delivered not only in specialized academic centers but also across diverse community settings.^16^

Safe administration requires careful monitoring for immune-mediated toxicities, particularly cytokine release syndrome (CRS) and immune effector cell–associated neurotoxicity syndrome (ICANS), which can occur during step-up dosing and early treatment cycles.^17,18^ In addition, long-term use predisposes patients to infection due to immunosuppression and hypogammaglobulinemia, further complicating management. Operational hurdles—including the need for hospitalization during dose escalation, infrastructure to provide urgent supportive care, and coordination between community and academic sites—add layers of complexity to widespread implementation^16,19^

Given these clinical and logistical considerations, clear guidance is required to inform best practices for the integration of BsAbs into routine myeloma care. This report synthesizes expert insights and evidence-based recommendations to support clinicians in patient selection, toxicity management, operational planning, and referral pathways. The goal is to enable safe, effective, and equitable delivery of BsAbs for patients with RRMM across both academic and community practice settings.

## Methods

This qualitative study used semi-structured interviews and thematic analysis to explore real-world implementation, toxicity management, and operational challenges associated with bispecific antibody (BsAb) therapy in relapsed/refractory multiple myeloma (RRMM). Participants were purposively selected to include clinicians with direct experience prescribing BsAbs or managing patients undergoing therapy.

### Data Collection

Interviews were conducted between November 2024 and May 2025 using a semi-structured interview guide. Interviews were conducted via videoconferencing, lasted approximately 45–60 minutes, and were audio-recorded with participant consent. Recordings were professionally transcribed verbatim for qualitative analysis.

### Participant Recruitment and Sampling

Participants were selected to include hematology/oncology clinicians from academic and community-based practices with experience managing patients receiving bispecific antibodies for relapsed/refractory multiple myeloma. Clinicians were identified through publication review and prior multiple myeloma educational programs and were invited to participate through direct outreach.

### Interview Guide

The interview guide focused on four core domains: (1) patient selection criteria and treatment sequencing, (2) toxicity monitoring and management strategies, including CRS and ICANS, (3) operational models for therapy delivery, such as inpatient versus outpatient ramp-up and academic–community referral structures, and (4) barriers and solutions related to long-term treatment access, equity, and sustainability. Interviews were semi-structured, allowing for open-ended discussion while ensuring consistent coverage of key topics. Transcripts were qualitatively analyzed to identify common themes, areas of consensus, and illustrative examples of best practices.

Sample interview questions included:

- “What is your experience managing patients with RRMM with bispecific antibodies?”
- “What is your preferred bispecific antibody to manage RRMM? What drives your selection?”
- “Is your practice equipped to treat patients with bispecific antibodies, or do you routinely refer patients to an academic center?”
- “How do patients know what adverse events to look for, when to contact someone, and who to contact?”
- “Who monitors for CRS (during step-up dosing and subsequent doses)? Has the monitoring and management of CRS been challenging at your practice?”

The full interview guide is provided in the supplemental materials.

## Results

### Theme 1: Perceived Clinical Efficacy and Role of Bispecific Antibodies in Late-Line RRMM

A consistent theme across interviews was the perception that BsAbs provide meaningful clinical benefit for patients with relapsed/refractory multiple myeloma who have exhausted other therapeutic options (Table 1). Clinicians described these agents as highly effective across multiple lines of therapy, including in triple-class refractory disease. The ability of these agents to induce remissions in patients who have limited alternatives underscores their perceived value among interviewed clinicians.

**Table 1.**
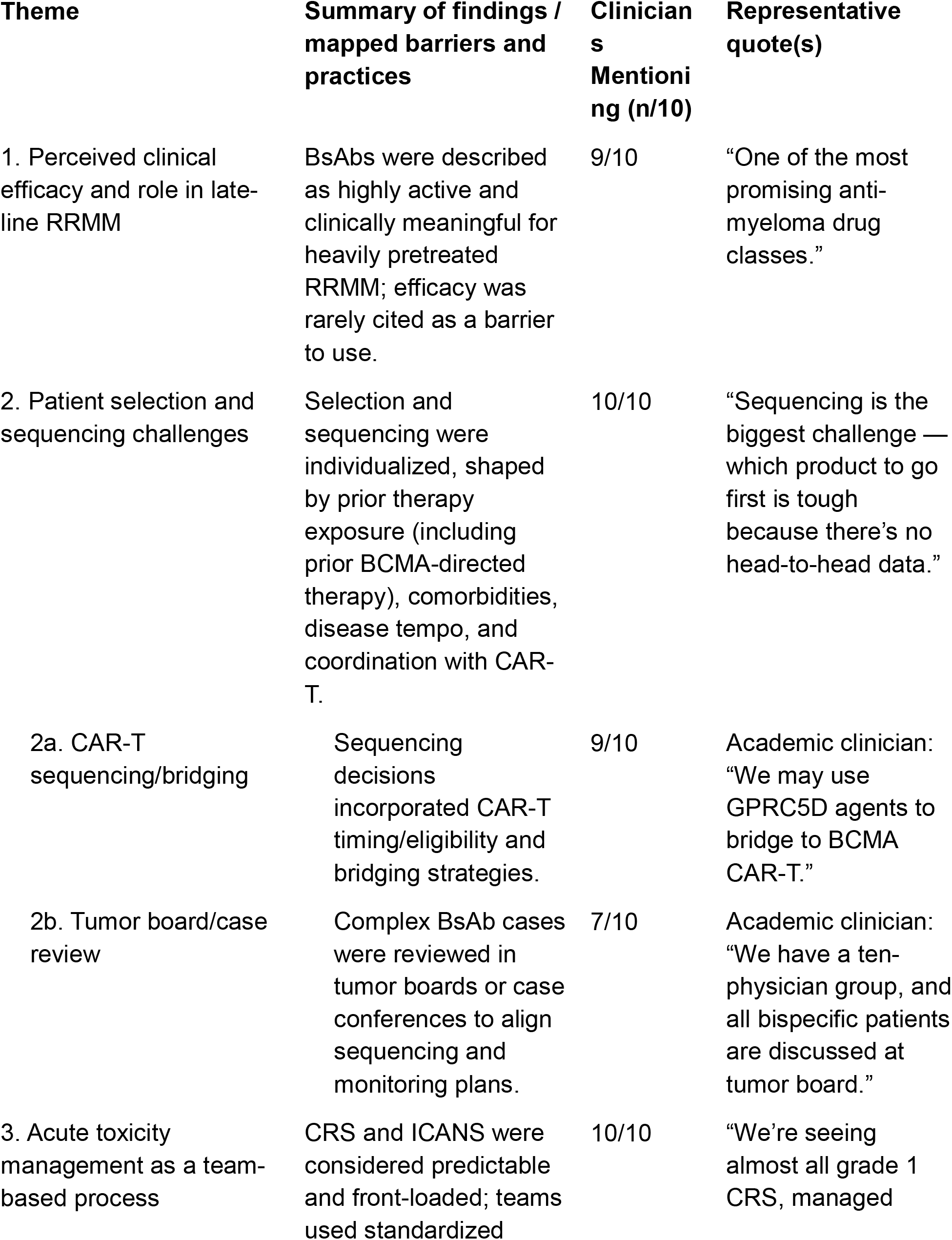
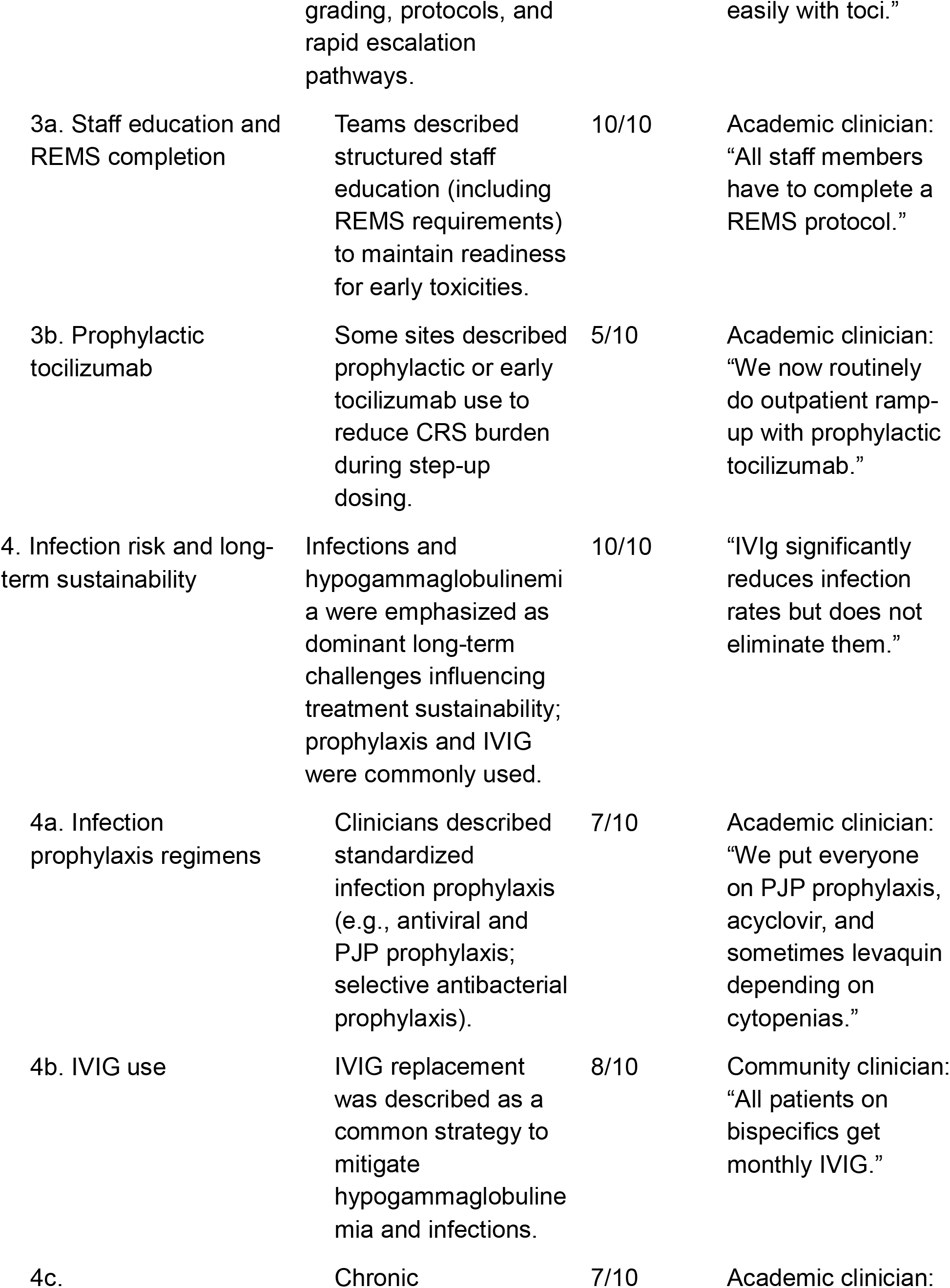
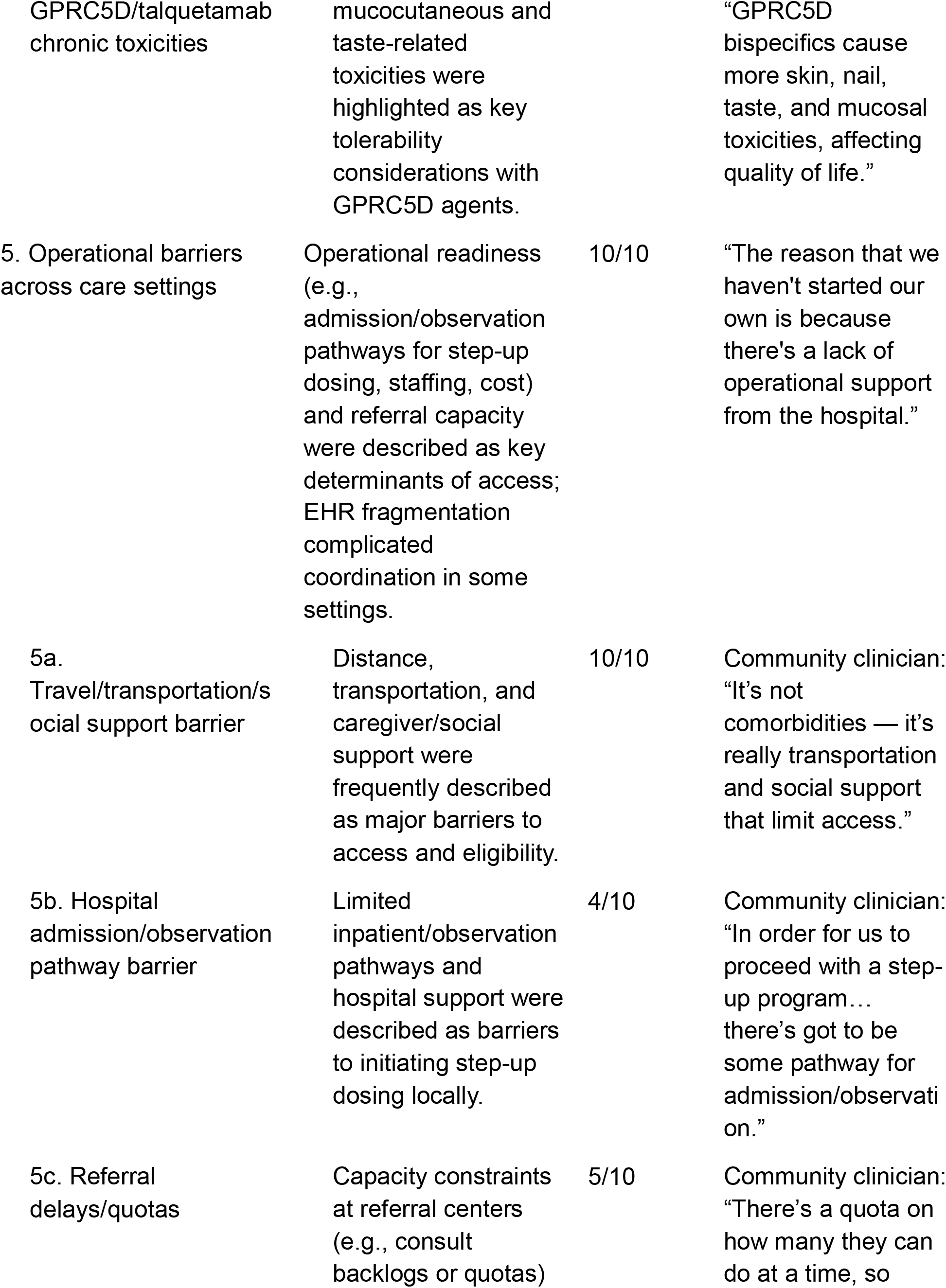
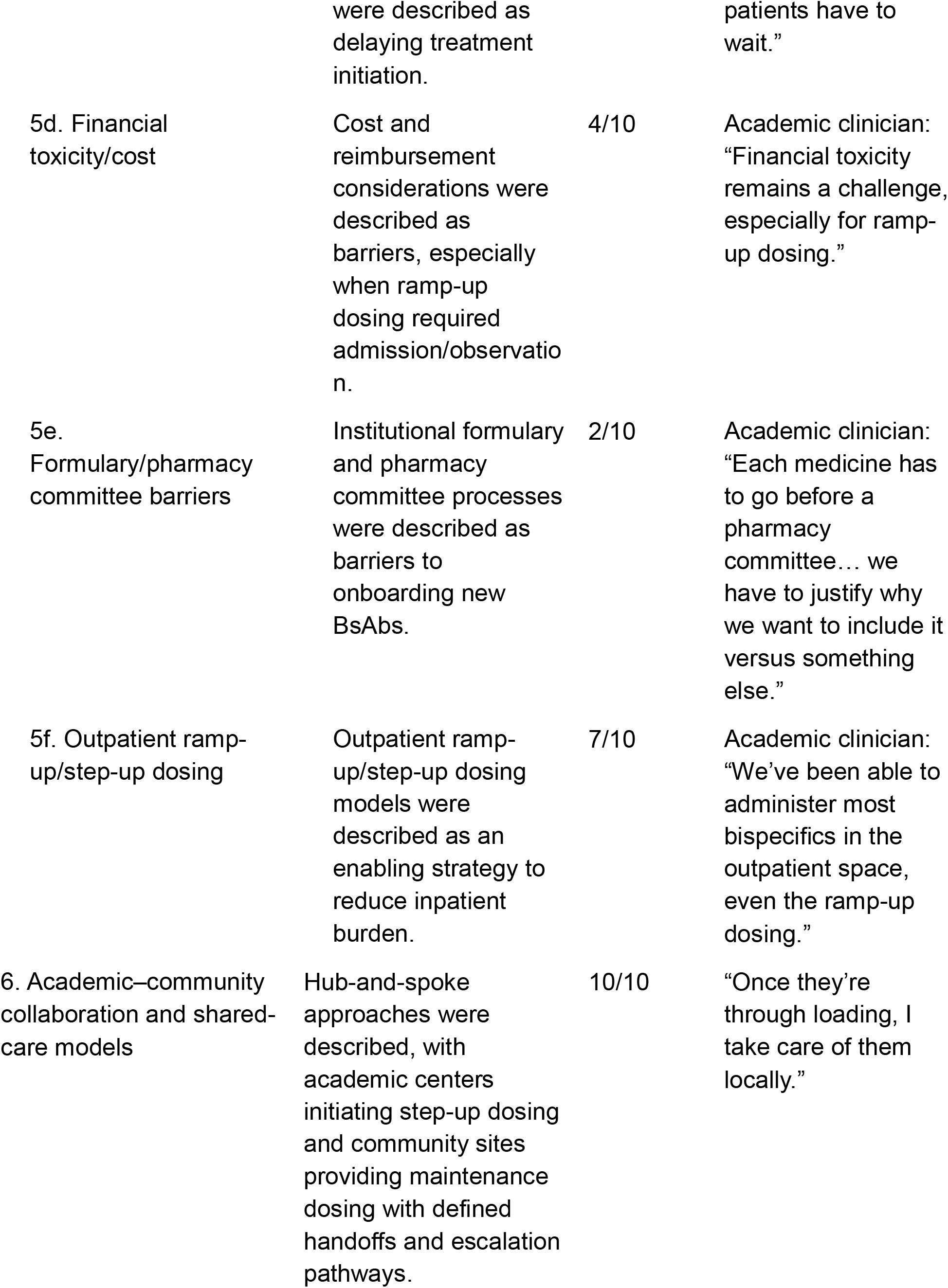
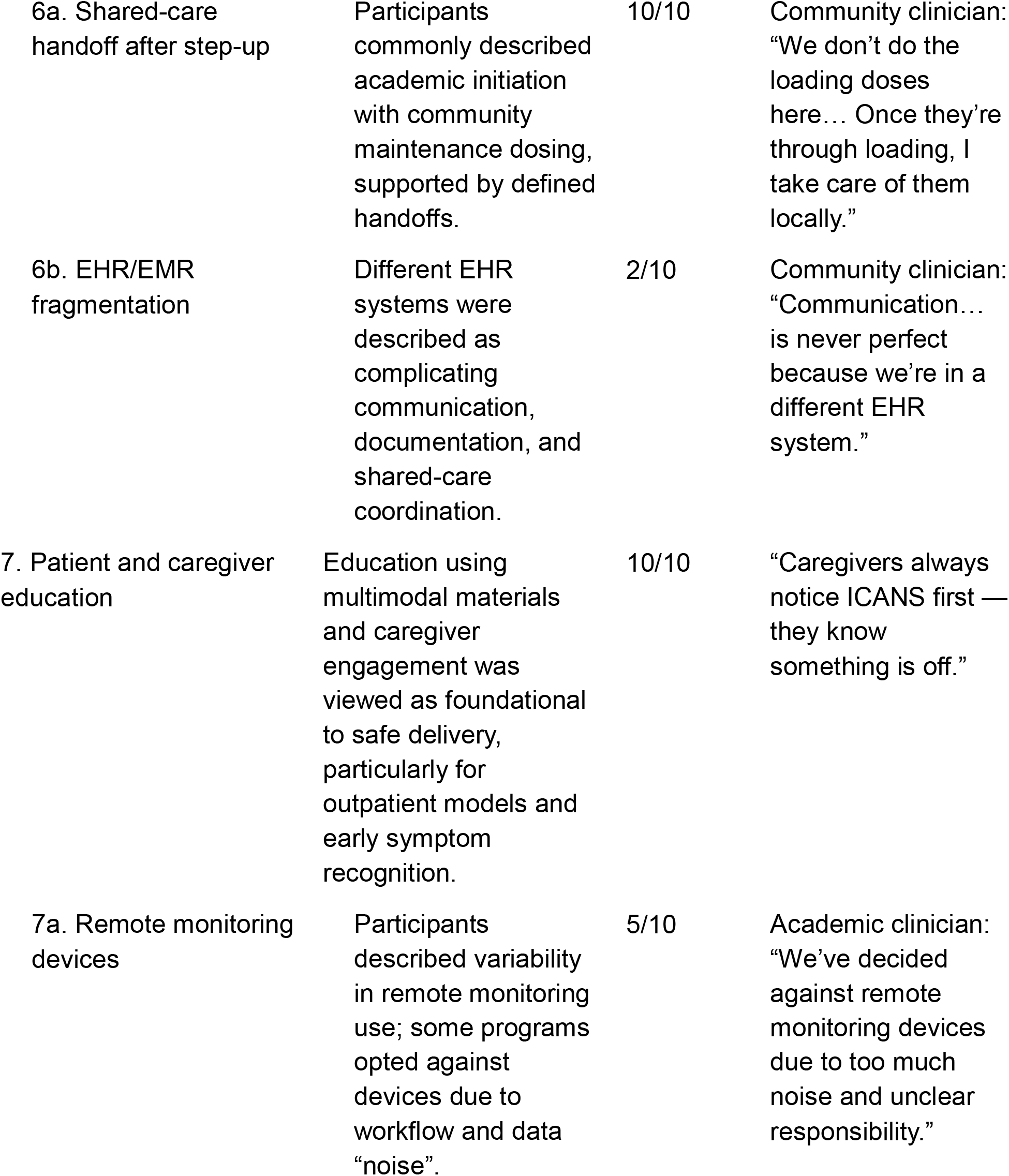
Summary of qualitative themes with representative quotes.

Illustrative quotes from participants included:

- “*Bispecific antibodies produce high response rates even in triple-class refractory patients*.”
- “*They stand out from many other options in the relapsed/refractory setting*.”
- “*Response can occur pretty quickly, and it deepens over time*.”
- “*This class is extremely promising and should be made accessible to as many patients as possible*.”

### Theme 2: Patient Selection and Sequencing as Ongoing Clinical Challenges

The absence of direct comparisons between B-cell maturation antigen (BCMA)- and G Protein-Coupled Receptor Class C Group 5 Member D (GPRC5D)-targeting agents has made therapeutic selection largely empirical. Clinicians described these decisions as highly individualized and frequently requiring multidisciplinary input, particularly in the absence of head-to-head comparative data and when coordinating BsAb therapy with CAR T-cell treatment (Table 1). Multidisciplinary case discussions, including tumor boards, were described as central forums for aligning treatment strategy across care teams. Treatment decisions often hinge on previous therapy exposures or anticipated transitions to other modalities, such as chimeric antigen receptor (CAR) T-cell therapy. Some centers use GPRC5D bispecific antibodies to stabilize disease before administering BCMA-targeted CAR T cells, reflecting a growing complexity in treatment sequencing and planning.^20^ This lack of comparative data presents one of the most significant clinical challenges today.

Illustrative quotes from participants included:

- “*We*’*re in this conundrum of trying to figure out how best to sequence therapies and identify the appropriate patient*.”
- “*We have a ten-physician group, and all BsAb patients are discussed at tumor board*.”
- “*There are patients who may have seen BCMA-targeted therapy beforehand, so if we know they*’*ve been exposed, we pivot to GPRC5D-targeting agents*.”
- “*A lot of selection is about prior therapy and the goal of using the bispecific*.”

### Theme 3: Acute Toxicity Management as a Predictable and Team-Based Process

Across interviews, clinicians described acute toxicities associated with BsAbs as expected and largely front-loaded, most commonly occurring during step-up dosing or early in treatment (Table 1). Cytokine release syndrome (CRS) was consistently described as the most frequently encountered acute toxicity, with participants noting that events were predominantly low grade and occurred early in the treatment course.

Clinicians described established institutional protocols and multidisciplinary team involvement as central to managing CRS when it occurred. Participants emphasized early recognition and timely intervention, including the use of tocilizumab, as common elements of their approach, reflecting increasing familiarity with BsAb-associated toxicities as clinical experience expanded.

In contrast to CRS, immune effector cell–associated neurotoxicity syndrome (ICANS) was described as less common but requiring heightened vigilance. Participants emphasized the importance of early detection strategies, including close neurologic monitoring and staff awareness, particularly during initial dosing periods. Ongoing team education and case review, including tumor boards, were described as mechanisms for reinforcing recognition of acute toxicities and maintaining operational readiness as experience with BsAbs evolved.

Illustrative quotes from participants included:

- “*CRS is expected, but it*’*s usually mild and very manageable if you intervene early*.”
- “*We*’*re seeing almost all grade 1 CRS, managed easily with tocilizumab*.”
- “*We host myeloma tumor boards and case reviews to keep education ongoing*.”

### Theme 4: Chronic Toxicities and Long-Term Treatment Sustainability

In contrast to acute toxicities, participants consistently described chronic toxicities as a key factor influencing the long-term sustainability of BsAb therapy (Table 1). Clinicians highlighted infection risk as a persistent concern over the course of treatment, noting that infections occurred even in the absence of neutropenia and often reflected cumulative immune dysfunction related to prior therapies and ongoing plasma cell depletion. While supportive strategies such as intravenous immunoglobulin (IVIG) replacement were commonly employed, participants emphasized that these measures reduced but did not eliminate the risk of infection.

In addition to infections, participants described cytopenias as a frequent and ongoing challenge requiring active management throughout treatment. Neutropenia was commonly cited as the most prevalent hematologic toxicity, with clinicians noting the use of growth factor support and close monitoring over time to allow continued therapy.

Participants also discussed toxicities specific to GPRC5D-targeted BsAbs, describing dermatologic, nail, oral, and taste-related adverse effects that could affect quality of life and nutritional status. These toxicities were characterized as distinct from those seen with BCMA-directed agents and were described as requiring patient counseling and supportive care rather than acute intervention, further contributing to considerations around long-term treatment tolerability.

Illustrative quotes from participants included:

- “*IVIg significantly reduces infection rates but does not eliminate them*.”
- “*Infectious disease is the most challenging to manage… even if they*’*re not neutropenic*.”
- “*Cytopenias are more likely in patients with high disease burden but tend to resolve over weeks to months*.”
- “*GPRC5D bispecifics cause more skin, nail, taste, and mucosal toxicities, affecting quality of life*.”

### Theme 5: Operational Barriers to BsAb Delivery Across Care Settings

Operational barriers were described as major determinants of whether BsAbs could be initiated and managed locally (Table 1). Community clinicians frequently described limited hospital support or pathways for step-up dosing (e.g., observation/admission, staffing and training requirements), and reliance on referral centers for initiation. Participants also described referral capacity constraints (e.g., consult backlogs or treatment quotas), formulary and pharmacy committee processes for new agents, and financial sustainability challenges for ramp-up dosing. Several participants highlighted communication barriers related to electronic health record (EHR) fragmentation, which complicated shared care and timely awareness of dosing and supportive care plans. This theme was raised by 10 of 10 participants.

Participants described substantial variability in where and how step-up dosing was delivered. Several academic programs reported moving portions of step-up dosing into outpatient infusion settings, supported by trained nursing teams, rapid response pathways, and ready access to tocilizumab. Others continued to rely primarily on inpatient admission or observation beds. Decisions about inpatient versus outpatient initiation were described as individualized and commonly informed by social support, travel distance, and baseline comorbidity.

Community clinicians described growing interest in developing local step-up dosing programs but noted that implementation required internal multidisciplinary planning (e.g., pharmacy, nursing, neurology, pulmonary/critical care) and recurring education to maintain team readiness. Administrative and operational details, such as inpatient drug cost attribution, local formulary status, and the ability to transmit records efficiently, were described as influencing whether initiation could occur locally or required referral.

Illustrative quotes from participants included:

- “*The reason that we haven*’*t started our own is because there*’*s a lack of operational support from the hospital*.”
- “*I have maybe a handful of contacts where I feel like communication flows well, but it*’*s never perfect because we*’*re in a different EHR system*.”
- “*Each medicine has to go before a pharmacy committee… we have to justify why we want to include it versus something else*.”
- “*We now routinely do outpatient ramp-up with prophylactic tocilizumab*.”

### Theme 6: Academic-Community Collaboration and Shared-Care Models

Clinicians described shared-care approaches in which academic centers initiate BsAbs (including step-up dosing and early-cycle monitoring) and community practices provide ongoing administration, monitoring, and supportive care once patients are clinically stable (Table 1). Participants emphasized that effective handoffs require clear documentation of dosing schedules and supportive care plans, direct clinician-to-clinician communication, and predefined escalation pathways back to the initiating center for acute toxicities or complex infections. This theme was raised by 10 of 10 participants.

Participants described multiple shared-care patterns, including hub-and-spoke models in which academic centers perform step-up dosing and early-cycle monitoring, followed by transition of maintenance dosing to local oncologists, as well as co-management models in which the initiating center remains involved through periodic reassessment, telemedicine touchpoints, or on-demand toxicity consultation. The content of handoffs commonly included dosing calendars, infection prophylaxis plans (including immunoglobulin monitoring and replacement thresholds), and criteria for dose holds, emergency evaluation, or re-referral.

Several community clinicians described that shared-care effectiveness depended on the timeliness and completeness of information exchange. Some participants reported receiving formal verbal and written signoffs and access to standardized protocols, whereas others described limited documentation or delayed communication, particularly when patients interacted with multiple referral centers or when EHR systems were not interoperable. Participants also noted variation across centers in supportive care practices (e.g., infection workup pathways and prophylaxis details), which could complicate co-management when patients transitioned between sites.

Illustrative quotes from participants included:

- “*We don*’*t do the loading doses here; we send them to [an urban hospital]. Once they*’*re through loading, I take care of them locally*.”
- “*If they go to other centers, I hear nothing. It*’*s very frustrating*.”
- “*Clear handoffs after cycle 1 and ongoing academic support help build confidence*.”
- “*Most of the patients return to their local provider after step-up dosing and are followed intermittently at the academic center*.”

### Theme 7: Patient and Caregiver Education as a Foundation for Safe Delivery

Participants described patient and caregiver education as foundational to safe BsAb delivery, particularly as more programs transition to outpatient step-up dosing and long-term outpatient maintenance (Table 1). Education strategies commonly included multimodal materials (printed handouts, booklets, videos), symptom checklists, and 24-hour contact numbers, reinforced through repeated teaching by clinicians, nursing teams, and pharmacy staff. Participants emphasized that caregivers play a central role in detecting early changes in cognition, fever, or functional status, and in supporting adherence to supportive care plans. This theme was raised by 10 of 10 participants.

Participants described education content as spanning both early immune-mediated toxicities (CRS and ICANS) and longer-term risks such as infections and cytopenias, with some clinicians also emphasizing counseling on chronic mucocutaneous and taste-related toxicities associated with GPRC5D-targeting therapies. Several sites described formal consent processes and the use of standardized materials such as wallet cards, written instructions, and escalation algorithms to clarify when patients should call the treating team versus seek emergency evaluation.

Approaches to home monitoring varied. Some sites reported providing basic home monitoring equipment (e.g., thermometer, blood pressure cuff, pulse oximeter) and conducting proactive outreach calls early in therapy, whereas others described avoiding wearable remote monitoring due to high signal volume and unclear responsibility for responding to alerts, instead relying on structured self-reporting and rapid access to the care team. Participants also noted that transportation barriers, limited health literacy, and language needs could influence the effectiveness of education and follow-up plans.

- “*Patients and caregivers receive at least two teaching sessions — one with a physician and one with nurses*.”
- “*We give patients an ICE score sheet, booklets on CRS/ICANS, and 24-hour contact numbers*.”
- “*I counsel patients extensively on logistics, hospital process, follow-up, and side effects*.”
- “*We train caregivers to recognize subtle signs like word-finding difficulty*.”
- “*We*’*ve decided against remote monitoring devices due to too much noise and unclear responsibility*.”

## Conclusion

BsAbs have demonstrated the ability to induce deep and durable responses in patients with RRMM, including those who have exhausted all standard therapeutic options.^12–15^ Their clinical promise underscores the importance of translating trial results into safe, practical, and equitable real-world use.^16,22^ In this qualitative study, clinician interviews (N=10) identified seven themes shaping real-world delivery across academic and community practice settings.

Successful integration of BsAbs into routine practice extends well beyond drug administration alone. Participants described acute toxicities (e.g., CRS and ICANS) as predictable and largely front-loaded during step-up dosing. These toxicities were manageable when programs use standardized, multidisciplinary workflows that included nursing-led monitoring, embedded grading and escalation algorithms, rapid access to tocilizumab and corticosteroids, and recurring team education. These findings align with emerging consensus guidance and published operational perspectives that emphasize protocolized monitoring, role clarity, and staff readiness for T-cell–engaging bispecific antibodies.^16,18,19,23–25^ Infection risk and long-term treatment sustainability were central concerns across interviews, with clinicians describing recurrent and sometimes atypical infections and treatment interruptions despite prophylaxis and immunoglobulin replacement. This theme is consistent with systematic and contemporary analyses reporting frequent and clinically meaningful infections and hypogammaglobulinemia during bispecific antibody therapy, particularly with BCMA-directed agents.^26–28^

To support safe expansion into outpatient and community settings, interviewees highlighted the need for operational models that strengthen academic–community partnerships and reduce access barriers (e.g., inpatient/observation capacity for step-up dosing, referral delays, travel and caregiver burden, financial sustainability, and electronic health record fragmentation; Figure 1). Hub-and-spoke shared-care approaches, where step-up dosing occurs at specialized centers and maintenance transitions to local practices, were described as enabling strategies when paired with structured handoffs, clear escalation pathways, and accessible academic back-up. Patient and caregiver education (multimodal teaching, consent and wallet cards, and 24/7 contact pathways) was viewed as foundational to outpatient safety and early symptom recognition, complementing published implementation roadmaps and best-practice recommendations for outpatient administration and coordination.^16,18,19,23–25,29^ By fostering collaboration across academic institutions, community providers, patients, and caregivers, the field can ensure that BsAbs are not only effective but also broadly accessible, paving the way toward more equitable outcomes in multiple myeloma care.

**Figure 1.**
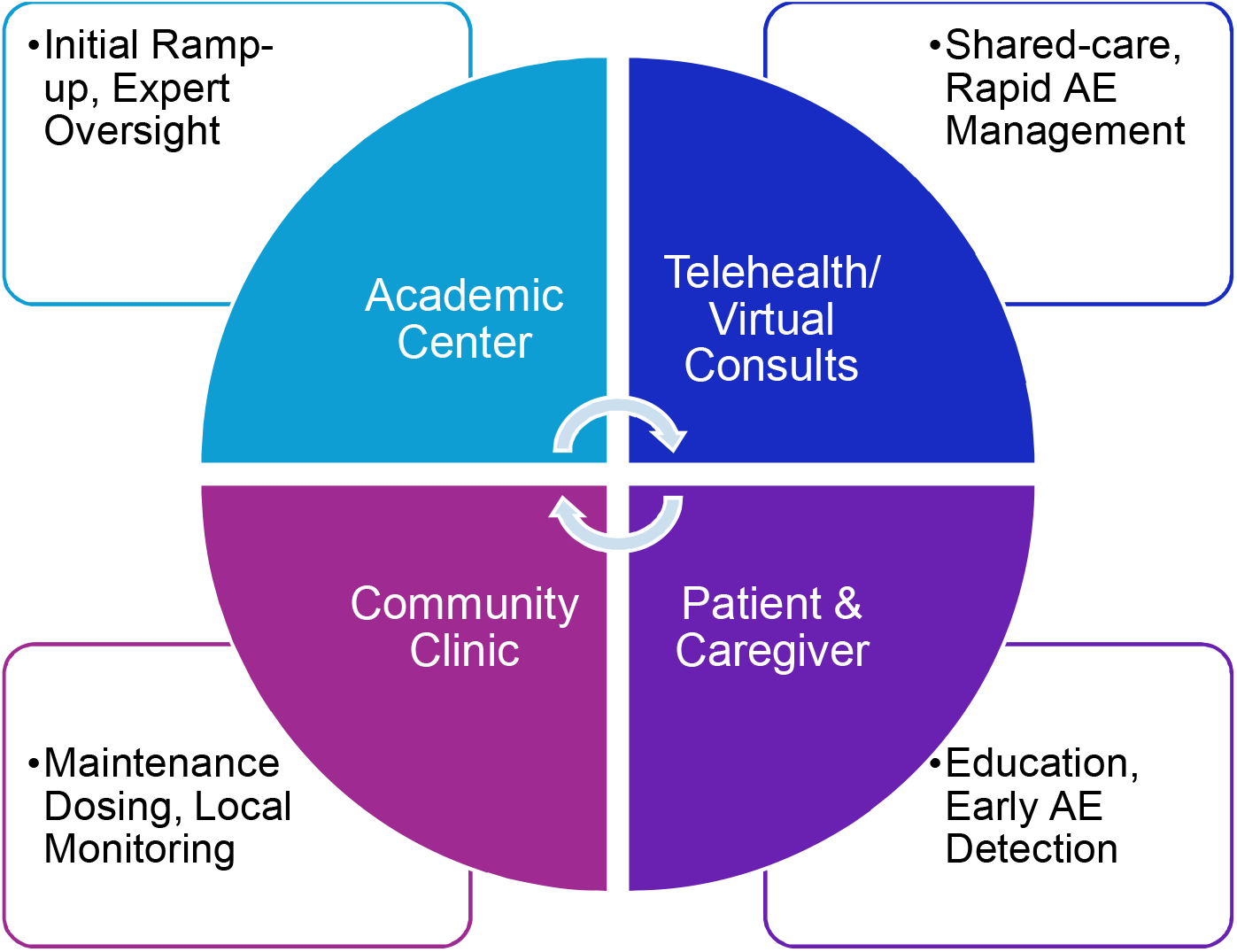
Academic-Community Collaboration Model for Bispecific Antibody Delivery.

## Data Availability

All data produced in the present study are available upon reasonable request to the authors

## Acknowledgements

Thank you to the academic and community hematologists/oncologists participating in this initiative and lending their insights and expertise. Thank you to the Medlive team involved in coordinating and implementing this educational design. This initiative would not be possible without support provided by an independent educational grant from Janssen Biotech.

## Supplemental Materials

### Full Interview Guide

#### Practice Questions

1. What is your experience managing patients with RRMM with bispecific antibodies (either in a clinical trial or commercially available)?
2. What is your preferred bispecific antibody to manage RRMM? What drives your selection? Is your preference based on patient population (ie, heavily pretreated ≥4 or 5 prior lines of therapy?
3. Are there specific patient populations that are more difficult to treat with bispecific antibodies?
4. How do you think the results of the investigational bispecific antibody linvoseltamab might impact your clinical practice?
5. Does your practice provide resources for racial or ethnic minority populations? (ie, translation services, etc?)
  a. Does your practice provide supportive care services to patients living in remote areas (ie, cover transportation, housing, and other expenses)?

#### Operationalization Questions

1. Is your practice equipped to treat patients with bispecific antibodies? Do you routinely refer patients to receive bispecific antibodies at an academic center?
2. Are there challenges with referrals? (ie, administrative, patient travel, communication with the academic center, etc)?
3. What are the top challenges to implementing novel bispecific antibodies into clinical practice?
4. Does your practice include discussions of bispecific antibodies in tumor boards (allows them to discuss each patient’s progress, therapeutic options, and treatment goals)? Does every patient go through a tumor board discussion? What are these discussions like? How do you decide whether a bispecific antibody is appropriate?
5. Is there education for all staff involved in the management of patients treated with bispecific antibodies? How often does your practice conduct team reviews of treatment plans? Which member of the care team is responsible for flagging AEs? Nurse or nurse practitioners?
6. How has your experience operationalizing bispecific antibodies been similar to other cellular therapies (ie, CAR T cell therapies?)
7. Which team member(s) are primarily responsible for follow-up care (treatment center or community oncologist)?
  a. Does your practice routinely use remote monitoring devices for your patients (ie, pulse, blood pressure, temperature, etc)? If so, how do patients report them?

#### Patient counseling on AEs and protocols for timely communication

1. What is your process for patient counseling for those initiating or switching a bispecific antibody?
2. Which member of the care team is primarily responsible for patient/caregiver treatment education and counseling?
3. **Probing question on communication**: How do patients know what adverse events to look for, when to contact someone, and who to contact? What is the follow-up process from there?

#### Management of CRS

1. Who monitors for CRS (during step-up dosing and subsequent doses)? Has the monitoring and management of CRS been challenging at your practice?
2. Are there any differences in patterns of onset, severity, and clinical manifestations among the various bispecific antibodies (BCMA-, GPRC5D-, or FcRH5-targeted) that you have noticed?
3. What labs do you routinely monitor for CRS? How frequently?
4. How is CRS managed?
  a. How do you manage CRS (low grade vs high grade CRS [grade 1 or 2 vs grade ≥3]?) Tocilizumab, dexamethasone, glucocorticoids, supplemental oxygen, single vasopresser, acetaminophen, others?
  b. What is the strategy after observing CRS (ie, dose modification, interruption, or discontinuation?)
  c. What type of prophylaxis are you most likely to recommend?
  d. Which care team member(s) are responsible for implementing supportive care and symptom management?
  e. How do you determine whether CRS will be managed by the community center or by the academic center?
  f. What role does nurse/nurse practitioners have in educating patients and their caregivers about managing expectations, toxicity monitoring and management?
5. Do you routinely consult with a specialist (ie, rheumatologist, intensivist, infectious disease specialist, immunologist, critical care physician) to assist with detection and management of CRS?
6. Does your practice have a standard operating procedure or guideline for CRS management?

#### Management of ICANS

1. Are there any differences in patterns of onset, severity, and clinical manifestations of ICANS among the bispecific antibodies (BCMA-, GPRC5D-, or FcRH5-targeted) that you have noticed?
2. Does your practice routinely utilize tools to monitor for ICANS (e.g., Immune Effector Cell Encephalopathy [ICE] score?
3. How do you engage family/caregivers with ICANS symptom recognition?
4. Does your practice have a standard operating procedure or guideline for ICANS management?
5. Which supportive measures do you currently use for ICANS? Tocilizumab, dexamethasone, levetiracetam?
  a. What is the strategy after observing ICANS (ie, dose modification, dose delay, interruption, discontinuation?)
  b. Approximately what percentage of your patients have required any of these supportive care measures?

#### Management of Infusion-related reactions (IRRs)

1. Where do your patients receive their treatment infusion? (in office/cancer center, infusion center, hospital, etc)
2. Do you believe the infusion staff is adequately prepared to manage IRRs? Who on staff can provide rapid management if needed? Is there communication between the community practice and academic center if an IRR occurs?
3. How are these currently managed?
  a. What prophylaxis/prophylactic regimens are you most likely to recommend? Topical corticosteroids and oral antihistamine?
    i. Probing question: Do you premedicate (dexamethasone or other agent) before the infusion date and the day of infusion to help prevent the IRR?
4. How are nurses and/or pharmacists educated on IRRs and management protocols for new therapies?
5. How often is the infusion schedule interrupted due to IRRs?

#### Management of Infections

1. Has the monitoring for infections been challenging at your practice?
2. Are there any differences in patterns of onset, severity, and clinical manifestations among the various bispecific antibodies?
3. How are infections currently managed?
  a. Who monitors for infections?
  b. At what point do you consult with an intensivist or infectious disease specialist?

#### Management of Hematologic AEs (ie, cytopenias, anemia, bleeding)

1. What types of hematologic AEs do you most commonly observe using teclistamab, talquetamab, erlantamab?
2. How do you approach management of hematologic AEs? Granulocyte colony-stimulating factor (G-CSF), intravenous immunoglobulin (IVIg)?
3. Is there hematologic toxicity that is more challenging than the others in terms of management?

